# Exploring Cancer in Colorado using a novel data platform: the ECCO experience

**DOI:** 10.64898/2026.02.03.26345489

**Authors:** Jan T Lowery, Faisal Alquaddoomi, Vince Rubinetti, Todd Burus, Cydney Jardine, Adam C Warren, Jake Walsh, Evelinn Borrayo, Sean Davis

## Abstract

**Purpose:** To create a publicly available, interactive data platform to visualize various data measures reflecting Colorado and its residents to support research and outreach efforts, specifically focusing on cancer burden and disparities throughout the state. This platform, named ECCO (Exploring Cancer in Colorado), aims to integrate diverse public data sources into a unified, user-friendly interface, accessible to researchers, community members, and outreach programs alike.

**Methods:** A multi-disciplinary team developed ECCO, leveraging public data sources like Cancer InFocus, State Cancer Profiles, and the Colorado Department of Public Health and Environment. The platform’s architecture employs a three-tiered web application model, utilizing a PostgreSQL database, a backend API built with FastAPI, and a Vue 3 frontend with an Open Layers map. Data is organized geographically at the county and/or census tract levels, categorized into measure categories (e.g., socio-demographics, cancer risk factors), and further filterable by demographic characteristics. An automated Extract-Transform-Load (ETL) data pipeline ensures regular updates of the data.

**Results:** The platform visualizes data such as socio-demographics, cancer risk factors, screening adherence, and cancer incidence and mortality rates. Additionally, ECCO incorporates location-specific data for cancer care facilities, health services, environmental exposures, and political boundaries. To date, ECCO has had 1.1K unique visitors and over 19K pageviews according to Google Analytics.

**Conclusion:** The ECCO platform provides a valuable tool for understanding and addressing cancer disparities in Colorado. By integrating diverse data sources and offering interactive visualization, ECCO enhances the ability of researchers, community members, and outreach programs to identify populations at risk, inform interventions, and support research priorities.

**Availability:** The application and code are available at https://coe-ecco.org/ and https://github.com/colorado-cancer-center/ecco.

**CONTENT SUMMARY:** *Key Objective:* This work sought to develop ECCO (Exploring Cancer in Colorado), an interactive, easily-accessible data platform designed to visualize and understand diverse cancer-related data measures reflective of Colorado and its residents.

*Knowledge generated:* ECCO integrates public data from sources like Cancer InFocus, State Cancer Profiles, and the Colorado Department of Public Health and Environment, visualizing measures such as socio-demographics, cancer risk factors, screening adherence, and cancer incidence and mortality rates at both county and census tract levels. The platform also incorporates location-specific data on cancer care facilities, health services, environmental exposures, and political boundaries.

## BACKGROUND

The University of Colorado Cancer Center (UCCC) is one of 73 NCI-designated comprehensive cancer centers across the U.S. and the only one in Colorado. The ‘catchment area’ for UCCC, which we define as the area and populations that we serve and intend to serve through our research, clinical services, and outreach activities, encompasses the entire state of Colorado. Understanding demographic factors, social determinants of health, environmental exposures, health behaviors, and the cancer burden as they vary across geographic areas and population subgroups is essential to providing equitable care and driving decisions about research priorities.

To this end, we sought to create a platform that would allow us to capture and visualize various data measures reflecting Colorado and our residents. At the onset, we envisioned building a platform that would serve multiple audiences, including UCCC researchers and staff, community members, and the UCCC Office of Community Outreach and Engagement (COE), which is charged with facilitating catchment-focused research and conducting outreach to reduce disparities in cancer burden in our state. Further, we wanted to build a platform that was broadly accessible, easy to use, and malleable, so that we could ingest new data to address emerging trends and needs of our catchment area. We were inspired in this effort by the work of several other NCI-designated cancer centers who had built or were building similar platforms, and who have graciously shared information about their processes and end-products.

We describe herein our interactive data platform called ECCO: Exploring Cancer in Colorado. We describe ECCO’s data content and sources, data model, and architecture that allows us to visualize various measures by multiple strata. We also provide several examples of inquiries that demonstrate the utility of ECCO in supporting research and in informing educational and outreach activities, as well as how we disseminate ECCO for broad reach.

## METHODS

### Team Organization and Requirements Gathering

To develop ECCO, a multidisciplinary team was formed, including experts in data science and informatics, software engineering, cancer epidemiology, population-based science, behavioral health, and disparities research. This team collaborated to define the project’s goals and scope. Initial steps involved a landscape analysis and vetting of existing data platforms to identify best practices and structural approaches. To understand end-user needs, the team developed detailed use cases and scenarios, which informed data set selection and platform functionality. These scenarios were crucial for prioritizing data collection, defining necessary platform features, and ensuring the platform’s usability for a diverse range of users, including researchers, community members, and outreach staff.Throughout the development process, regular meetings and feedback loops were implemented to revise and refine requirements based on user feedback and evolving project goals. Presentations to the Cancer Center membership, COE team, and Cancer Advisory Council, as well as national meetings, resulted in additional high-level feedback and ensured alignment with larger Catchment initiatives.

### Data Sources and Prioritization

ECCO utilizes a diverse array of public data sources,listed at https://coe-ecco.org/sources. The data model is primarily inspired by Cancer InFocus (CIF), providing data on socio-demographics, housing, economic measures, environmental exposures, and health behaviors through its monthly release^1^. Cancer incidence and mortality data are sourced from State Cancer Profiles (SCP), also updated monthly, stratified by sex, cancer stage, age, and race^2^. Beyond county and census tract level data, ECCO integrates location-specific data from CIF’s “US facilities and providers” table, focusing on cancer treatment centers and sites of public health interest. Additional Colorado-specific data sources include the Colorado Cancer Resource Map (CCRM)^3^, radon testing, HPV vaccination rates from the Colorado Department of Public Health and Environment (CDPHE), and political district information from official Colorado and US government websites. Data sources were prioritized based on their relevance to cancer burden and disparities in Colorado, data availability, and accessibility for regular updates.

### Data Model and Updates

ECCO’s data model organizes values by geographic entity (county and census tract) and categorizes them into measure categories (e.g., sociodemographics). Measures are specific values associated with these geographic entities. The model includes “factors” for data subset filtering, such as sex, cancer stage, age, and race for SCP data. Locations of specific entities like cancer care facilities and political boundaries are also integrated. Primary datasets are updated monthly via an automated Extract-Transform-Load (ETL) pipeline, which acquires, processes, and loads data into a PostgreSQL database. This pipeline ensures data consistency and currency.

### Platform Architecture and Technology

ECCO is a three-tier web application: a PostgreSQL database, a backend API built with FastAPI, and a Vue 3 frontend with an Open Layers map. This architecture enhances scalability by allowing the backend to serve numerous requests efficiently. The frontend provides a user-friendly interface with selectable data categories, measures, and locations displayed on an interactive map. Users select the category, measures, factors, and locations they want to show with simple dropdowns. Measures are plotted on the map as colored counties or tracts. Locations are plotted at specific points as markers of a particular color (and shape, for accessibility) or enclosed areas of a particular dashed outline. Contextual tooltip functionality provides additional details and/or links to “additional details” pages. While the interface is simple, it offers advanced customization options. All software tools used are open-source, including PostGIS for geographic data, Alembic for schema migrations, uvicorn for backend processes, TypeScript for robust frontend development, and Snakemake for reproducible workflows. All ECCO code is available on GitHub at https://github.com/colorado-cancer-center/ecco/.

## RESULTS

We launched ECCO in August 2024, making it accessible directly at https://coe-ecco.org/. ECCO serves as a key tool for characterizing our catchment area, pinpointing populations experiencing cancer disparities, and supporting research endeavors within our region. Currently, there are 174,577 records stored in 22 database tables, all accessible via the ECCO UI. We track 140 distinct measures within these 22 tables (Supplemental Material, Table 1). The following highlights some key results and insights derived from the platform.

### Population Demographics

ECCO’s visualizations provide detailed demographic breakdowns, including age groups, educational levels, race and ethnicity, and various socioeconomic factors. For example, **Figure 1** reveals a higher percentage of Hispanic residents in many rural counties of southeastern and northeastern Colorado. This data is critical, as Hispanics, being the largest and one of the fastest-growing minority groups in the state, face significant cancer care and outcome disparities. This insight helps identify key geographic areas for targeted study recruitment and community outreach.

**Figure 1:**
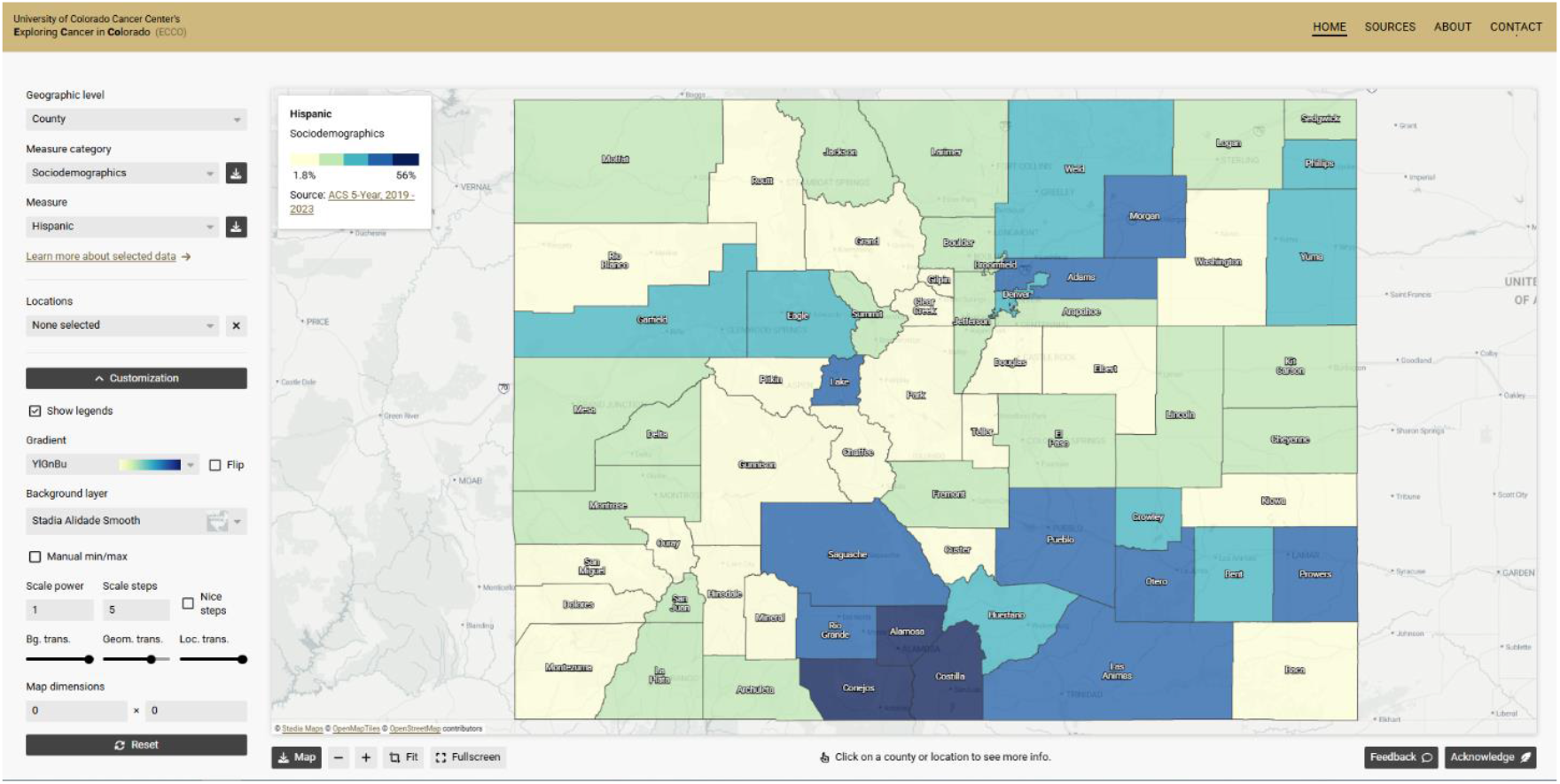
Demographics: Percent of Colorado population that is Hispanic by county

### Cancer Risk Factors

The platform includes data on various personal and environmental risk factors for cancer. **Figure 2** demonstrates the variation in current smoking rates across counties, ranging from 8.5% to 19%, and differences in potential radon exposure by displaying the percent of homes tested with radon levels above the EPA-recommended threshold. These visualizations can be used to inform and target educational campaigns and evidence-based interventions, particularly in areas with higher risk levels. For instance, counties with higher smoking rates may benefit from targeted smoking cessation programs.

**Figure 2:**
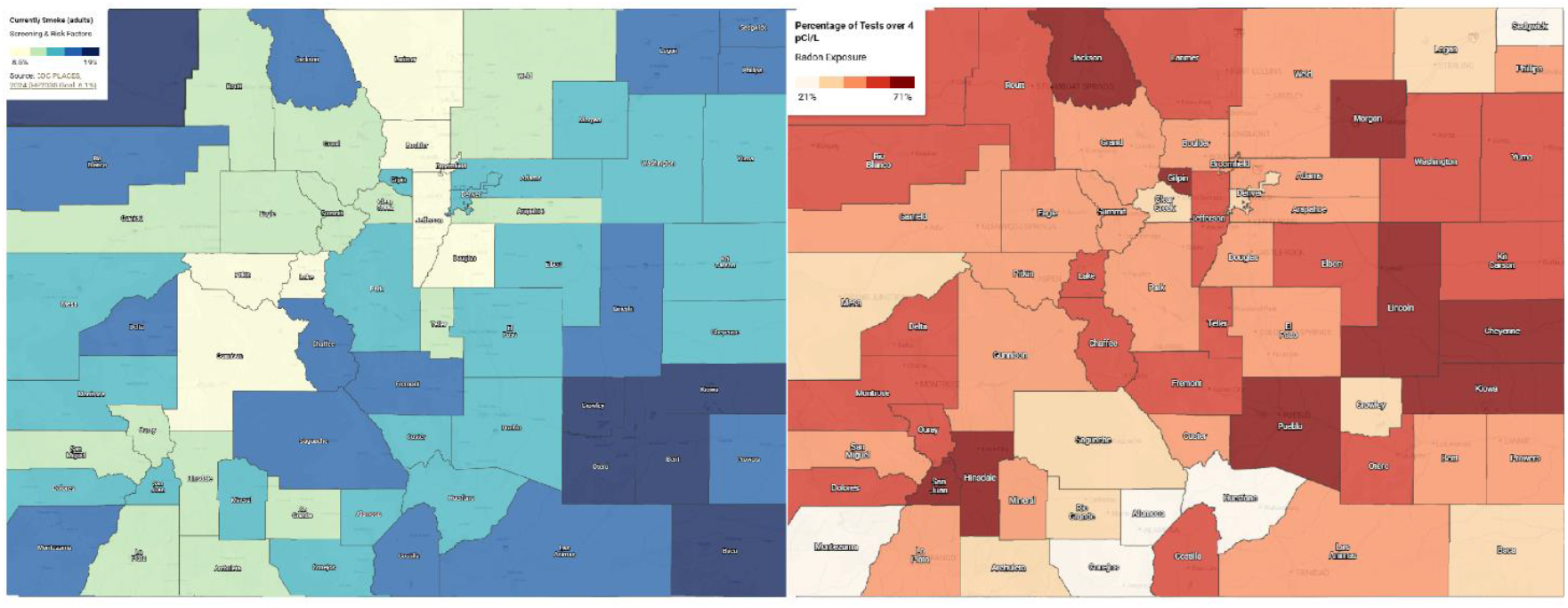
Risk Factors: Tobacco and Radon

### Cancer Screening

ECCO visualizes adherence rates for recommended cancer screenings, such as mammography, colorectal, and cervical cancer. **Figure 3** illustrates mammography adherence among women aged 50-74, with lighter-colored counties indicating lower rates. Overlaying locations of mammography screening centers (red dots) reveals disparities in access, particularly in rural and mountain counties, which could explain the lower screening rates observed in these areas. This suggests a need to improve access to screening in these regions.

**Figure 3:**
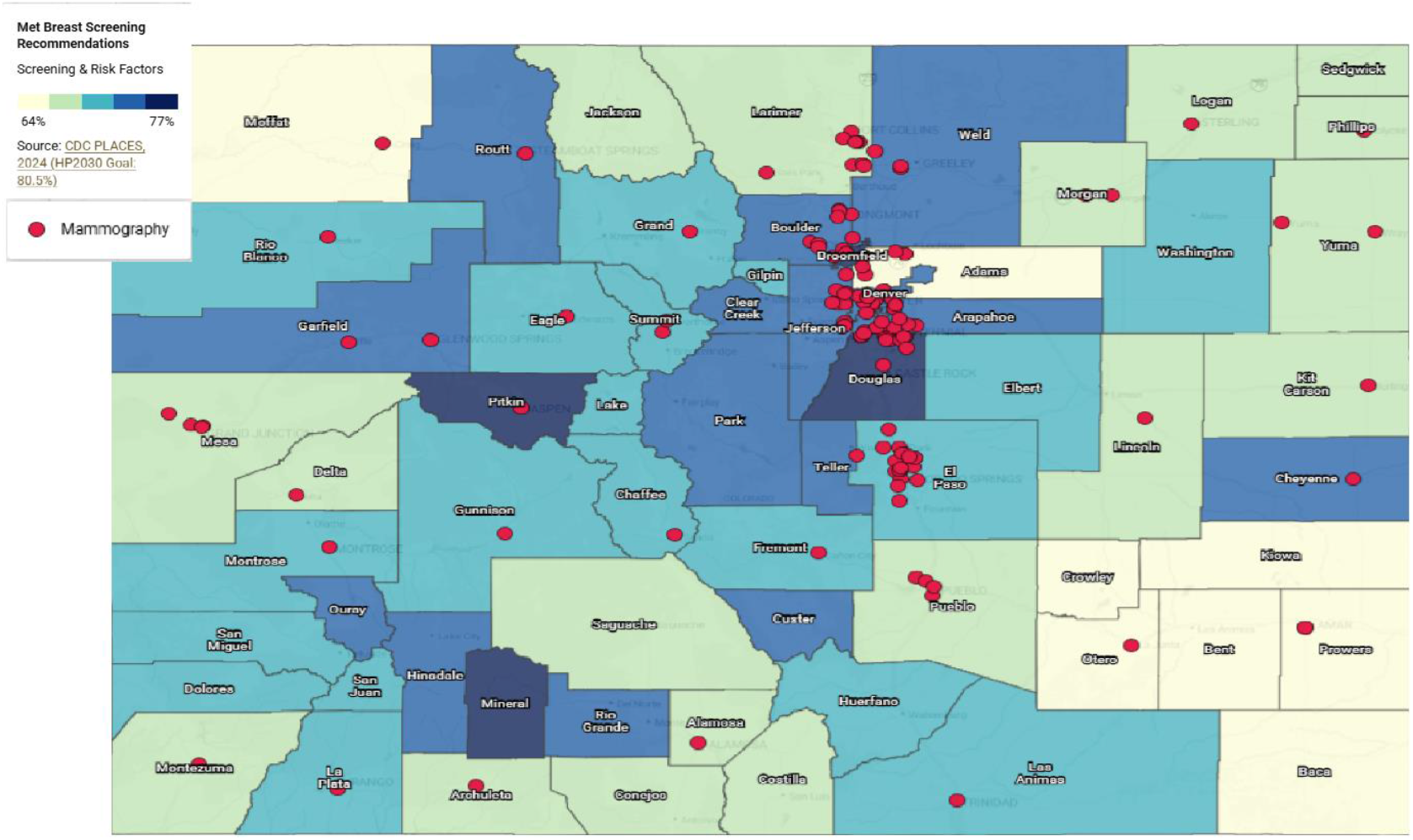
Cancer Screening: Breast cancer screening with mammography locations.

### Cancer Burden

ECCO displays cancer incidence and mortality rates, averaged over five years, along with five-year trend data by county. **Figure 4** shows colorectal cancer incidence rates, with darker areas indicating higher rates. When a user clicks on a county, ECCO provides the specific rate and the average annual count of new cases. Additionally, users can filter incidence data by sex, stage, race, and age group, allowing for nuanced investigations of cancer burden. These trends are critical to help identify areas where public health outreach and prevention may be most effective.

**Figure 4:**
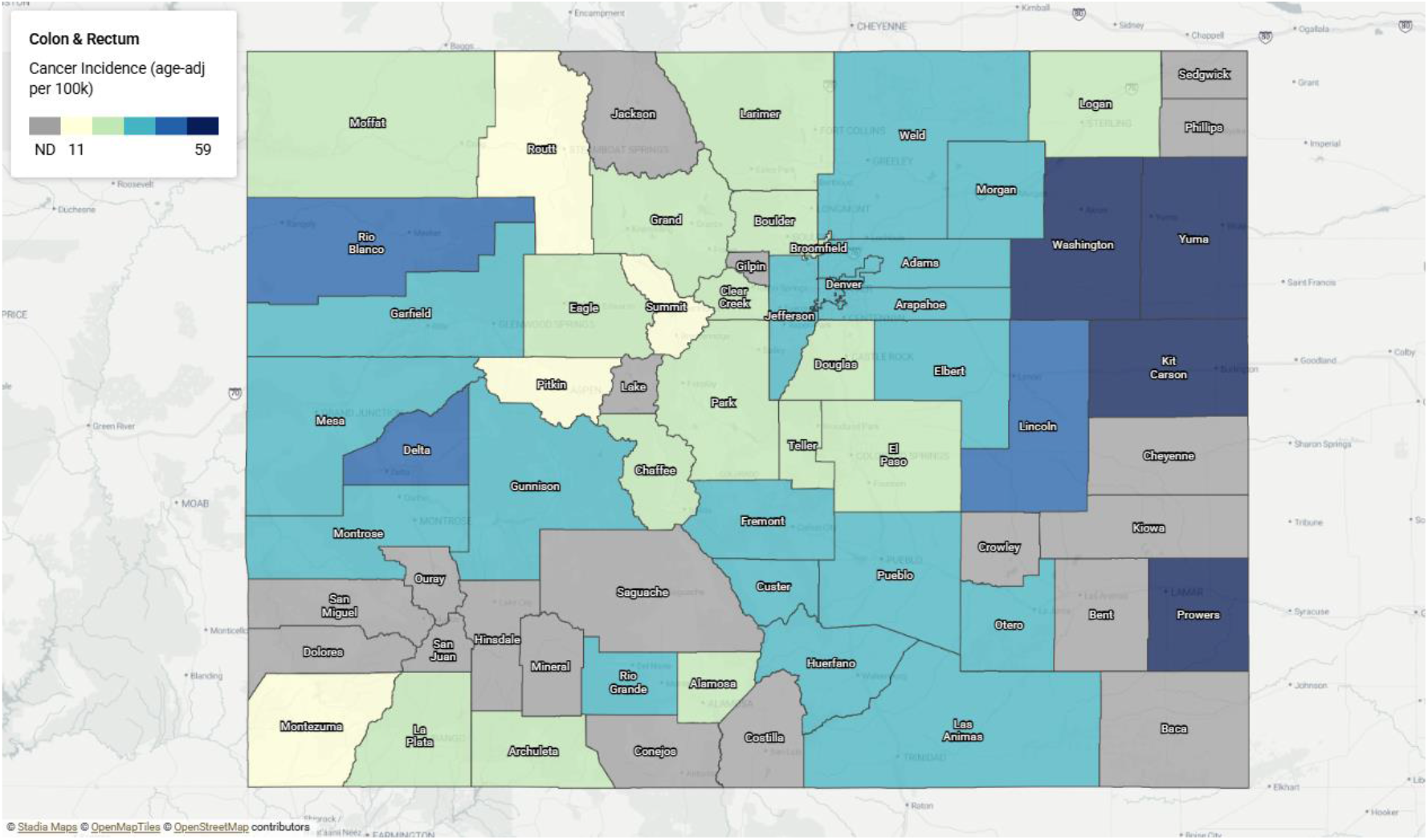
Trends in Incidence: CRC

### Added Functionality and Data Access

In addition to visualization, ECCO allows users to download data in tabular format for use in grants, publications, and presentations. County-specific data summaries, as seen in **Figure 5** for Pueblo County, offer comprehensive snapshots with graphs and tables. Furthermore, overlaying legislative districts and contact information for representatives facilitates informed educational messaging and policy briefs. Finally, COE activity locations are mapped, aiding in evaluating outreach effectiveness. These multiple functionalities, combined with ease of data download, extend ECCO’s value to researchers and public health professionals.

**Figure 5:**
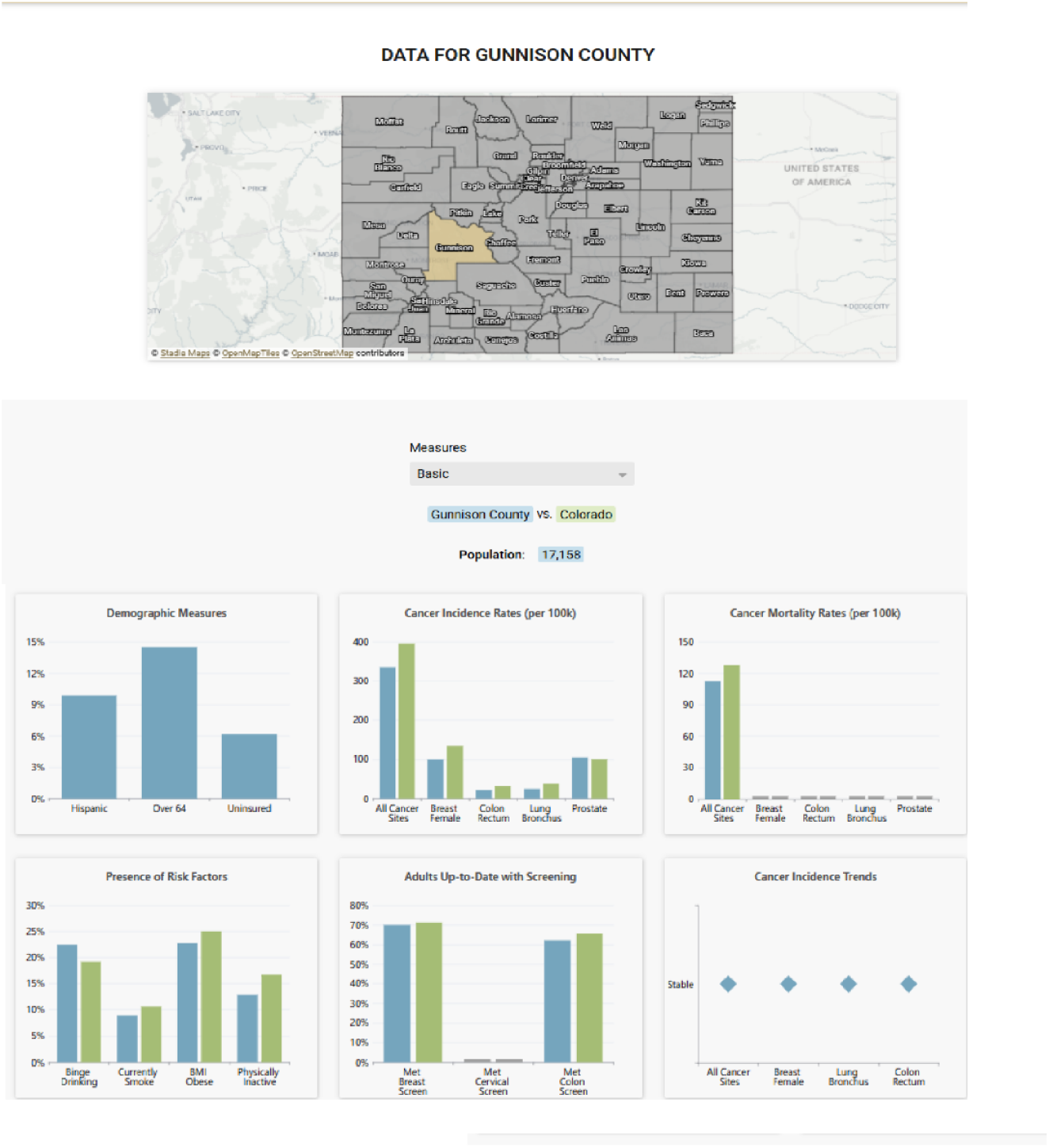
County Snapshot: Pueblo

### ECCO Uptake and Engagement

ECCO has garnered 1.2K unique visitors and over 19K pageviews based on Google Analytics data. Dissemination efforts have included UCCC blogs ([https://news.cuanschutz.edu/cancer-center/ecco-interactive-cancer-mapping] (https://news.cuanschutz.edu/cancer-center/ecco-interactive-cancer-mapping) ), presentations to the Community Advisory Council and other internal and external groups, and demonstrations at various community events. Furthermore, ECCO has been designated as a community resource in the upcoming Colorado state cancer plan. This broad engagement indicates the value of ECCO to different users.

## DISCUSSION

The ECCO platform stands as a valuable asset for the University of Colorado Cancer Center (UCCC) and the communities it serves. It effectively addresses the need to characterize Colorado’s diverse population and cancer-related data in a user-friendly, intuitive web-based platform. ECCO has greatly enhanced our ability to identify areas and population subgroups experiencing disparities in cancer risk, outcomes, and access to care. Moreover, it empowers UCCC researchers to explore and justify new research ideas that directly respond to the needs of our catchment area.

Software and Data Engineering for Flexibility and Scalability: One of ECCO’s key strengths lies in its robust software and data engineering approaches, which ensure a flexible and scalable product. The fully automated data ingest system eliminates the need for manual acquisition and integration of data from upstream sources, saving significant time and resources. The review process implemented for each monthly release allows administrators to catch and rectify any issues arising from changes in upstream data schemas. By adopting a “long table” schema at the measure category level, we avoid needing schema changes for newly introduced measures. Additionally, the schema migration system ensures that all schema changes are meticulously tracked over time, maintaining data integrity and reproducibility. ECCO’s three-tiered web application architecture further enhances scalability, enabling efficient handling of numerous small requests from a large number of clients, unlike session-based architectures.

Successful Application of Multidisciplinary Team Science: ECCO represents a successful application of multidisciplinary team science collaboration, bridging departments and disciplines across the UCCC and extending to the broader cancer informatics community through the CARDS (Catchment Area Research and Data Science) working group. This national network of cancer centers has provided invaluable knowledge sharing, solutions, and shared challenges, which significantly aided ECCO’s development. Specifically, our partnership with the data science team at the Markey Cancer Center (University of Kentucky) was critical to the project’s success. This collaborative approach facilitated the integration of diverse expertise, from data science and software engineering to cancer epidemiology and community outreach, leading to a well-rounded and effective platform.

### Ongoing Growth and Maintenance

ECCO’s value will continue to grow with ongoing expansion and refinement. We have plans to add new data sources to address emerging concerns in our catchment area, such as youth and adult vaping prevalence, lung cancer screening uptake, and UV light intensity data. Feedback from our Community Advisory Council (CAC), community partners, and UCCC research teams is continuously sought to improve content and functionality. A new partnership with the Multidisciplinary Center on Aging has spurred consideration of adding data sources relevant to cancer and aging research, including chronic conditions that often co-occur with cancer in older adults. These additions and refinements will ensure ECCO remains a relevant and dynamic tool.

### Clear Vision and Strong Implementation for Success

The development and success of ECCO underscore the importance of having a clear vision supported by strong implementation. The initial vision of creating an accessible, user-friendly platform for multiple audiences, including researchers, community members, and outreach staff, guided all subsequent decisions. However, it was the meticulous attention to implementation—from the initial landscape analysis and use case development to the technical architecture and data pipelines—that truly brought the vision to life. This careful execution, combined with continuous evaluation and adaptation, has been instrumental in creating a valuable and impactful resource for the UCCC and its catchment area.

### Challenges and Lessons Learned

Several challenges were encountered, and valuable lessons were learned along the way. Due to ECCO’s public-facing nature, some sensitive data cannot be displayed. Additionally, small numbers in rural counties may lead to data suppression to protect privacy. These limitations have necessitated careful consideration of data display and dissemination strategies. The team also had to balance stakeholder input and resource availability to evaluate new features and data requests, and discuss extending ECCO’s scope to ensure that proposed new work could be performed successfully, efficiently, and effectively.

## Conclusion

ECCO provides a critical tool for understanding and addressing cancer risk,incidence, mortality, and disparities in Colorado. By leveraging robust software and data engineering, fostering interdisciplinary collaboration, and maintaining a commitment to ongoing development, ECCO has enhanced and will continue to positively impact the UCCC’s capacity for research, outreach, and community engagement.

## Data Availability

All data are available publically on our ECCO platform at www.coe-ecco.org

https://github.com/colorado-cancer-center/ecco

## ACKNOWLEDGEMENTS

We would like to acknowledge the CARDS (Catchment Area Catchment Area Research and Data Science) working group. This national network of cancer centers has provided invaluable knowledge sharing, solutions, and shared challenges, which significantly aided ECCO’s development.

