## Supplemental Table 1 for "Exploring Cancer in Colorado using a novel data platform: the ECCO experience"

**SUPPLEMENTAL MATERIAL**

Table 1: Currently available ECCO data categories and examples of measures in each.

| Measure Category | Level | Measures/Sites |
|---|---|---|
| Cancer disparities index | County | Breast Cancer Index, Colorectal Cancer Index, Head and Neck Cancer Index, Lung Cancer Index |
| Disparities | County + Tract | Economic Segregation, Gender Pay Gap, Gini Coefficient, Lack_English_Prof, Living Below Poverty (Black), Living Below Poverty (Hispanic), Racial Economic Segregation, Racial Segregation, Uninsured (Black), Uninsured Children, Uninsured (Hispanic) |
| Economy | County + Tract | Annual Labor Force Participation Rate, Annual Unemployment Rate, Below Poverty, Household Income, Insurance Coverage, Medicaid Enrollment, Monthly Unemployment Rate, Received Public Assistance, Uninsured |
| Environment | County | LILATracts_Vehicle, pct5G_35_3, pct5G_7_1, pctBB_1000_100, pctBB_100_20 |
| Environment | Tract | Diesel PM, Drinking Water Noncompliance, Hazardous Waste Proximity, Lead Paint, Nitrogen Dioxide, Ozone, PM25, RMP Proximity, Superfund Proximity, Toxics Release to Air, Traffic Proximity, Underground Storage Tanks, Water Discharge |
| Food desert | Tract | LILATracts_Vehicle |
| Housing and transportation | County + Tract | Crowded Housing, Lack Complete Plumbing, Median Gross Rent, Median Home Value, Median Monthly Mortgage, Mobile Homes, Multi-Unit Structures, No Home Broadband, No Vehicle, Owner Occupied Housing, Rent Burden (40% Income), Single Parent Household, Vacancy Rate |

| | | |
|---|---|---|
| HPV | County | Up-To-Date Percent |
| Radon | County + Tract | NTests, NTestsover4, PctOver4 |
| Risk factors and screening | County + Tract | Asthma, Bad_Health, Binge_Drinking, BMI_Obese, BP_Medicine, Cancer_Prevalence, CHD, Cognitive_Disability, COPD, Currently_Smoke, Depression, Diabetes_DX, Food_Insecure, Had_Stroke, Hearing_Disability, High_BP, High_Cholesterol, Housing_Insecure, Independent_Living_Disability, Lacked_Reliable_Transportation, Lacked_Social_Emotional_Support, Met_Breast_Screen, Met_Colon_Screen, Mobility_Disability, No_Teeth, Physically_Inactive, Poor_Mental, Poor_Physical, Recent_Checkup, Recent_Dentist, Selfcare_Disability, Sleep_Debt, Socially_Isolated, Vision_Disability |
| Cancer mortality | County | All Cancer Sites, Bladder, Brain & ONS, Breast (Female), Cervix, Childhood (Ages <20, All Sites), Colon & Rectum, Esophagus, Kidney & Renal Pelvis, Leukemia, Liver & Bile Duct, Lung & Bronchus, Melanoma of the Skin, Non-Hodgkin Lymphoma, Oral Cavity & Pharynx, Ovary, Pancreas, Prostate, Stomach, Thyroid, Uterus (Corpus & Uterus, NOS) |
| Cancer incidence | County | All Cancer Sites, Bladder, Brain & ONS, Breast (Female), Breast (Female in situ), Cervix, Childhood (Ages <15, All Sites), Childhood (Ages <20, All Sites), Colon & Rectum, Esophagus, Kidney & Renal Pelvis, Leukemia, Liver & Bile Duct, Lung & Bronchus, Melanoma of the Skin, Non-Hodgkin Lymphoma, Oral Cavity & Pharynx, Ovary, Pancreas, Prostate, Stomach, Thyroid, Uterus (Corpus & Uterus, NOS) |
| Sociodemographics | County + Tract | 18 to 64, Advanced Degree, AIAN, Asian, Below 9th grade, Black, College, High School, Hispanic, NHOPI, |

| | | |
|---|---|---|
| | | Other_Races, Over 64, Total, Under 18, White |
| Cancer Incidence | State | All Cancer Sites, Bladder, Brain & ONS, Breast (Female), Breast (Female in situ), Cervix, Childhood (Ages <15, All Sites), Childhood (Ages <20, All Sites), Colon & Rectum, Esophagus, Kidney & Renal Pelvis, Leukemia, Liver & Bile Duct, Lung & Bronchus, Melanoma of the Skin, Non-Hodgkin Lymphoma, Oral Cavity & Pharynx, Ovary, Pancreas, Prostate, Stomach, Thyroid, Uterus (Corpus & Uterus, NOS) |
| Cancer Mortality | State | All Cancer Sites, Bladder, Brain & ONS, Breast (Female), Cervix, Childhood (Ages <15, All Sites), Childhood (Ages <20, All Sites), Colon & Rectum, Esophagus, Kidney & Renal Pelvis, Leukemia, Liver & Bile Duct, Lung & Bronchus, Melanoma of the Skin, Non-Hodgkin Lymphoma, Oral Cavity & Pharynx, Ovary, Pancreas, Prostate, Stomach, Thyroid, Uterus (Corpus & Uterus, NOS) |
| Sociodemographics | State | Binge Drink, Currently Smoke (adults), Met Breast Screening Recommendations, Met Cervical Screening Recommendations, Met Colorectal Screening Recommendations, Obese (BMI over 30), Percentage of Tests over 4 pCi/L, Physically Inactive |
